# Causation and prevention in epidemiology: assumptions, derivations, and measures old and new

**DOI:** 10.1101/2024.12.20.24319429

**Authors:** Robert Allard

## Abstract

Epidemiologic measures quantifying the causative or the preventive effect of a particular agent with respect to a given disease are frequently used, but the set of assumptions on which they rest, and the consequences of these assumptions, are not widely understood. We present a rigorous derivation of these measures from the sufficient-causes model of disease occurrence and from the definition of causation as the bringing forward of the occurrence time of an event. This exercise brings out the fact that an understanding of the assumptions underpinning all measures of effect, and of the extent to which they may or may not be met, is necessary to their prudent interpretation. We also introduce a new measure, discarding 1) the sufficient-causes model and 2) the assumption that the agent can only be either causative or preventive, relative to a given disease, but not both. Some may consider this more acceptable than having to decide, on slim or no evidence, that the agent has only one kind of effect on the disease. In any case, I submit that epidemiology should eventually discard the concept of causation, as has been done in some other basic sciences, and replace it with the adequate modeling of disease-producing processes, in individuals and populations.

## Introduction

The existence and nature of causation have been discussed since at least Greek antiquity. Currently, there is in epidemiology a broad consensus that an agent is causative with respect to a disease (or other outcome, such as death) in an individual, if exposure to the agent brings forward the time of occurrence of the disease in the individual, compared to what it would have been had the individual not been exposed to the agent (1-8). This may mean that the disease will now occur during this individual’s lifetime, instead of not occurring. Conversely, an agent is considered preventive if it delays the time of occurrence of the disease. This may mean delaying it until after the individual has died, that is, the disease never occurring in this individual.

This communication presents, starting out from an earlier publication of the author (9), a theoretical framework for deriving quantitative measures of the causative or preventive effect(s) of an agent. It explains some commonly used measures and the assumptions on which they rest, and provides an alternative measure, not currently used but which has advantages over the usual ones.

## Methods

The sufficient-causes model of disease causation postulates that a disease occurs in an individual exactly at the moment when one sufficient cause of this disease gets completed, that is, when all the component causes that make up this sufficient cause are present in this individual (1). A causative agent is a component cause of at least one sufficient cause. Conversely, an agent preventive of a disease is one whose absence is required in at least one sufficient cause of the disease, that is, its presence blocks the completion of this sufficient cause. Thus, in theory the same agent can be both causative and preventive of a disease in the same individual, if the other component causes of at least one sufficient cause of each type are present in the individual. There may also be other sufficient causes of the same disease that are unaffected by the presence or absence of the agent. For any disease, there may be many sufficient causes of each of the three types.

An equivalent approach for our present purposes is to think of the sufficient cause as a process or mechanism with distinct steps (the component causes), when the last of which is completed disease immediately occurs. For brevity, in the rest of this text the unqualified word *cause* will refer to a sufficient cause.

## Results

In order to operationalize this approach, let us assume that we have a population of individuals each of whom potentially incorporates all three types of causes leading to the disease of interest. For simplicity and without loss of generality, we will further assume that only one cause of each of the three types is potentially present in any individual (see Appendix 1 for the justification). We will also assume that there are no competing risks, including that of death, making it impossible for the individuals to develop the disease after a certain time.

We assume that each cause has a given completion time in each individual, when the last component cause falls into place. We will represent these completion times, measured from T = 0, the beginning of the follow up period for an individual at risk for the disease, by t_1_, t_0_ and t_**·**_, respectively for the cause requiring the presence of the agent, the one requiring its absence and the one unaffected by it. In an exposed individual t_0_ is counterfactual, and in an unexposed one t_1_ is.

At follow-up time T, in an exposed individual, if t_1_ < (T, t_0_ and t_·_), the disease has occurred, caused by the presence of the (causative) agent completing a sufficient cause. In an unexposed individual, if t_0_ < (T, t_1_ and t_·_), the disease has occurred, caused by the absence of the (preventive) agent not blocking a sufficient cause. In any individual, if t_·_ < (T, t_1_ and t_0_), the disease has occurred, unaffected by the presence or absence of the agent.

Over all at-risk individuals in a given population, each cause is completed in individuals at an incidence rate (number of completions over person-time at risk), expressed as a function of time t, of i_1_(t), i_0_(t) and i_·_(t), respectively (see Appendix 2).

To account for the three causes operating simultaneously, we postulate the existence of a joint probability density function JPDF(i_1_, i_0_, i_·_) of the completion times of the three causes. As mentioned earlier, in this theoretical reasoning, we will ignore the existence of competing risks that would prevent the causes from ever reaching completion, but we will bring up this issue again in the discussion.

Following our definition of causation, the probability that the cause requiring the presence of the agent is going to get completed before the other two causes is given by

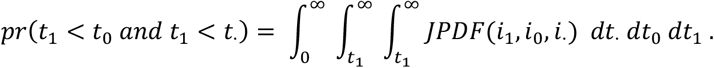

The inner integral indicates that t. > t_1_, the middle one that t_0_ > t_1_ and the outer one that t_1_ can be any time after the start of follow-up. The probabilities of the other causes getting completed first are defined similarly, *mutatis mutandis*. Appendix 3 solves the integral, under the further assumptions that 1) the completion times of the causes are independent of each other (the *independence of occurrence times* assumption), so that their completion rates are additive, and 2) these rates are constant over time (the *constancy of incidence rates* assumption), to make the integral solvable. The result is

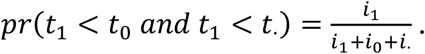

Similarly, one gets

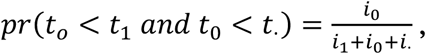

and

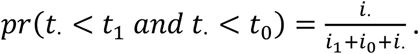

Since an individual can only be either exposed or unexposed to the agent of interest, it is impossible to estimate all three quantities i_1_, i_0_ and i_·_. In practice, one estimates the disease incidence rate among the exposed, I_E_, or the cumulative risk R_E_, and the corresponding quantities among the unexposed, I_U_ and R_U_. (Uppercase Is will represent observable rates and lowercase i’s derived ones.) If I_E_ > I_U_ (or R_E_ > R_U_), one assumes that i_0_ = 0, that is, that the agent is never preventive, so that there exist only causes that either require its presence as a component or that are unaffected by it (the *homogeneity of effect* assumption). Under this assumption and the other two, we have I_E_ = i_1_ + i_·_ and I_U_ = i_·_ and therefore

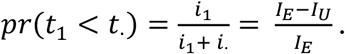

This quantity is often referred to as the *attributable risk (percent)*, as the *etiologic fraction* (EF), or as the *assigned share*, and by other names. If one defines the hazard ratio as HR = I_E_/I_U,_ one gets EF = (HR – I)/HR. As approximations, the risk ratio RR = R_E_/R_U_ or the odds ratio OR = R_E_(1-R_U_) / (1-R_E_)R_U_ are sometimes substituted for HR in the expression for the EF.

One usually interprets the EF as representing the proportion of cases (occurring among susceptible persons exposed to the agent of interest) that is, in some sense, attributable to, or caused by, the agent. It must not be confused with the proportion of cases attributable to the agent among the whole susceptible population, exposed and unexposed, called the *population EF*.

Conversely, if I_U_ > I_E_ (or R_U_ > R_E_), one assumes that i_1_ = 0, that is, that the agent is never causative, so that there exist only causes that are either blocked or that are unaffected by its presence. Then we have we have I_E_ = i_·_ and I_U_ = i_0_ + i_·_ and

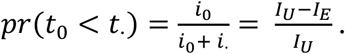

This quantity is often referred to as the *prevented, preventive* or *preventable fraction* (PF) and by other names. Using HR, we get PF = 1 – HR. Again, HR is sometimes replaced by RR or OR.

The *homogeneity of effect* assumption is problematic, as we never know all the causes of which an agent is a component cause. There are agents known to have both causative and preventive effects on the same disease; for instance, mammography both prevents and causes breast cancer, the preventive effect being of course much larger than the causative one.

There is an alternative, which seems never to have been considered so far: that there are no causes in which the agent play no role, that is, that i_·_ = 0. Then we have I_E_ = i_1_ and I_U_ = i_0_ and

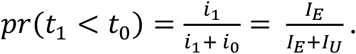

We have proposed the name *causal fraction* (CF) for this quantity (9). By discarding the *homogeneity of effect* assumption, the CF is able to quantify the net effect of both causative and preventive causes. In other words, it is unconditional on whether the agent is causative or preventive. If both types of causes have no effect whatsoever on disease occurrence or have equivalent effects, I_U_ = I_E_ and CF = ½. If causative causes predominate CF > ½ and if preventive causes predominate CF < ½.

Using the same unconditional approach, we can also get

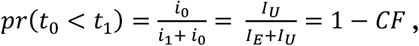

which confirms that there is no need for a separate measure of preventive effects, since it would only be the complement of CF.

A very important property of the CF is that one can derive it directly from observed disease incidence rates, without recourse to the sufficient-causes model (Appendix 4). Thus, CF can be interpreted simply as the probability that the disease will occur sooner under exposure to the agent than under non-exposure.

The CF can also be derived from a constant hazard ratio HR without knowing the values of the terms I_E_ and I_U_ of the ratio, whether these rates are constant (Appendix 5), or variable over time (Appendix 6). Thus

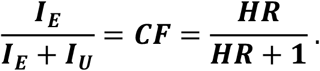

## Discussion

The CF does not require recourse to the sufficient-causes model or to the *homogeneity of effect* assumption. It does require the *constancy of incidence rates* assumption and that of *independence of occurrence times* t_1_ and t_0_ if it is to take the simple form I_E_/(I_E_+I_U_).

Rejecting the *homogeneity of effect* assumption and assuming instead that all causes leading to a disease are more or less affected by the presence or absence of an agent is apparently novel. However, nothing in this assumption prevents some of theses causes from being of negligible practical importance, after being amalgamated with all the other causes that operate in the same direction, either causative or preventive (Appendix 1).

Randomization best ensures that one can attribute any observed difference between I_E_ and I_U_ to the presence or absence of the agent, that is, that no confounding is present. In other words, randomization is our best means of achieving *group exchangeability*, meaning that the distribution of disease occurrence times in the unexposed comparison group is the distribution that we would have observed in the exposed group, had it been unexposed. Thus, randomized controlled trials (RCTs) are the context in which the CF (or any other measure of association) most credibly lends itself to a causal interpretation.

One can often present the results of a RCT as two survival curves from the time of randomization to the time of experiencing the event or of censoring, one curve among those exposed to the agent or treatment of interest and the other among those unexposed. Under *group exchangeability*, a method is available for estimating the minimum and maximum values of the CF compatible with the two distributions (see reference 9 and Appendix 7). Some general results of this method are: a) If all occurrence times under exposure are shorter than the shortest occurrence time under non-exposure, then CF=1. Conversely, if all occurrence times under exposure are longer than the longest occurrence time under non-exposure, then CF=0. b) If the distribution of occurrence times is exactly the same under exposure as under non-exposure, then, surprisingly, as N→∞, the bounds tend toward 0≤CF≤1 (9). c) For situations where the two distributions overlap partially, one can estimate the minCF and maxCF compatible with both distributions (reference 9 and appendix 7). d) If *individual exchangeability* could be achieved, that is, if each exposed subject could be associated with one unexposed subject whose disease occurrence time truly represented his/her counterfactual disease occurrence time under non-exposure, then CF would equal the proportion of exposed individuals who experience disease sooner than their unexposed counterpart (9) and we would have minCF = CF = maxCF.

Irrespective of randomization, the EF, PF and CF can all be estimated in a survival analysis of groups of exposed and unexposed subjects, using the HR estimated by proportional hazards regression (10), particularly if the ln[-lnS(t)] plots are parallel, and preferably taking competing risks into account (11). This still requires the assumptions of *independence of occurrence times*, but not that of *constancy of incidence rates* (Appendix 6) which gets replaced by the less stringent assumption of *constancy of the hazard ratio*.

Finally, EF, PF and CF are all vulnerable to random and systematic errors affecting the measurement of disease occurrence times or the estimation of incidence rates.

## Conclusion

Even supposing that we have met all assumptions required for the CF to be valid, there remains at least one concern: how to measure the *combined* effects of all agents on the disease. The question is all the more important that the *homogeneity of effect* assumption implies that every agent have some effect, however small, on every disease.

Modelling, such as by using directed acyclic graphs, is one way to represent the relationships between variables, some of them treated as causes, others as effects, others as both. Useful representations of causal webs are rarely simple. Considerable information and judgment are needed to select the variables and relationships that deserve to be included without cluttering the model with unimportant elements, especially in a situation where everything is considered *a priori* relevant. When one seeks *individual exchangeability*, such as by nearest neighbor, propensity or disease risk score matching, modelling of this kind is required to select the best control(s) for any given case. Modelling is also required in counterfactual reasoning, to help decide what divergences from reality, from among the set of all that one can imagine, are compatible with each other, and in the creation of propensity scores, where it helps select the initial set of potential matching variables and avoid matching for intermediate variables (“overmatching”). Ideally, these models should not be generic statistical ones, but should rather take into account the specific properties of every component (“nodes” or “vertices”) and of their interrelationships (“edges” or “arcs”).

Astrophysicists are able to predict with exquisite precision the position and velocity of all the permanent components of the inner solar system, from the distant past to the distant future, using a relatively small number of measurements. The same ability to predict the evolution of a system exists in many other branches of physics. It rests on having a model of the system that is practically definitive. Most importantly here, the concept of causation, which is in any case intellectually problematic and difficult to operationalize, plays no role in these models, in spite of their describing perfectly the relationships between all components of the system, or rather *because* they do so. Once a system is thoroughly described by a model, causality evaporates.

I submit that, as for astrophysics (*pace* quantum theory), epidemiology deals with phenomena that are essentially continuous, although historically, for reasons of habit and convenience, it has segmented the processes it studies into phases, such as genetic predisposition, later risk factors, disease, treatment, complications and death. Depending on the researchers’ focus, they label some phases as causes and others as effects, without there being, in fact, any essential difference between the two categories. To avoid these categories while retaining the ability to indicate the direction in which a continuous process is moving, we could replace the nouns “cause” and “effect” with *antecedent (phase)* and *subsequent* (*phase*, rather than *consequent*, because of its causal connotation).

Although the CF is interpretable in terms of causation, to a non-causal way of thinking its also being interpretable purely in terms of exchangeability is a step in the right direction. A name for the CF that does not refer to causation would be helpful in this regard. *Subsequent fraction*, or *ensuing fraction*, might fit the bill, but I welcome suggestions.

## Data Availability

All data used are hypothetical and included in the article.

## Appendices

Appendices 1 to 6 present, for readers interested in the most explicit mathematical explanations, some generally accepted epidemiologic concepts necessary for following the main text, and elaborate derivations of the measures of effect discussed in it. Each appendix is as independent of the others as possible, at the expense of some repetition.

Appendix 7 illustrates the calculation of minCF and maxCF from distributions of disease occurrence times under exposure and non-exposure.

### 1 Independence of the occurrence times of events implies additivity of their occurrence rates

Assume that there are n sufficient causes, all requiring the presence of the agent, that lead to the disease of interest. Let us call R the probability that any one of them, or several ones, are completed by time T, and r_x_ the probability that sufficient cause x among them will have been completed by time T. The corresponding probabilities of non-completion are (1-R) and (1-r_x_), respectively. For the disease not to occur by time T, no cause must get completed by time T. Under the *independence of occurrence times* assumption, by the well-known principle that the joint probability of independent events is the product of their individual probabilities, the overall probability of disease non-occurrence is the product of the cause-specific probabilities of non-completion:

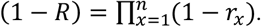

If the overall *probability* of completion R is related to the constant *rate* of completion I of any cause, and if each cause-specific probability r_x_ is related to rate i_x_, by the usual relationship (Appendix 2) we have

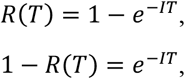

where R(T) is the cumulative risk of disease at time T. Similarly for the separate causes:

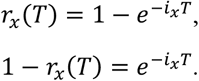

The first equation now becomes

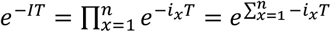

Therefore

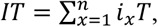

and finally

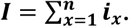

Thus, if we assume the independence of the occurrence times of the sufficient causes, the rate of first completion of any cause acting in a given direction is the sum of the rates of completion of the separate causes acting in that direction. In other words, independence of occurrence times of causes implies additivity of their rates of occurrence. We can therefore treat all the separate sufficient causes of the same type as one cause, whose completion rate is the sum of the completion rates of the individual causes.

Obviously, the same reasoning applies to the causes that require the absence of the agent and to those that are unaffected by it, so that we can reason adequately about the indicators using only 3 sufficient causes. One can also carry out this reasoning without the *constancy of incidence rates* assumption, the exponents being definite integrals (Appendix 2).

### 2 Relationship between rate and risk

Since a full explanation does not seem to be available online, we reproduce it here, from the one textbook that presents it (1, pp. 29-31). It begins by defining the instantaneous time-dependent incidence rate I(t) at time t, in a time-dependent at-risk population P(t) at time t, in the absence of competing risks. Given the absence of competing risks, this incidence rate I(t), is the instantaneous decrease in the size of the at-risk population at that time, −*dP*(*t*), over the population-time at the same time, *P*(*t*) *dt* :

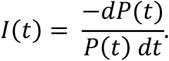

Therefore

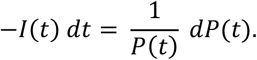

Since 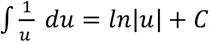, u being any function, P(t) in this case, integrating both sides of the equation gives:

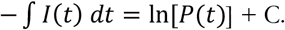

The definite integral from time 0 to time T is

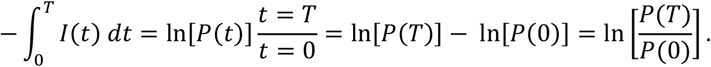

Since *e*^ln *x*^ = *x*, exponentiating the first and last sides gives

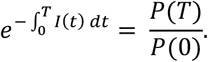

Now, the cumulative risk of disease up to time T is defined as

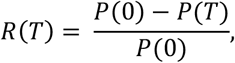

that is, R(T) is the number [P(0) – P(T)] of individuals now missing from the at-risk population over the initial at-risk population P(0); again, in the absence of competing risks they must be missing because they contracted the disease. Therefore

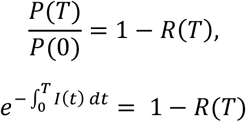

and finally

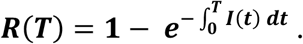

At T = 0, R(T) = 1, and as T → ∞, R(T) → 1. This latter relationship may not hold for diseases of childhood that will never occur in an individual if they have not occurred by a certain age, creating “immortal time”.

If the rate I is constant over time, since ∫ *I dt* = *It*, the formula for R(T) becomes

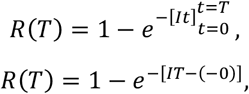

and finally

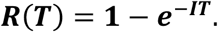

An elaborate and recent discussion of the relationship between risk and rate is available (12).

### 3 Solving the basic triple integral

The probability that the sufficient cause involving the agent will occur before the other two sufficient causes is given by

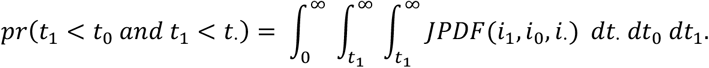

Under the *independence of incidence times* assumption, the joint probability density function JPDF is the product of the probability density functions for each of the three causes, so that

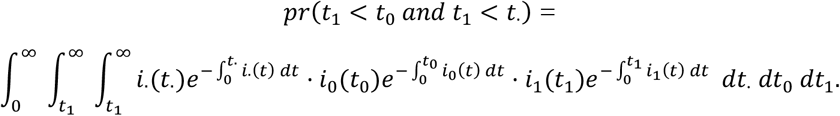

Under the assumption that i_1_, i_0_ and i_·_ are constant instead being of functions of time (the *constancy of incidence rates* assumption), the above equation becomes

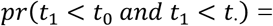

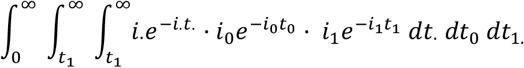

Integrating successively from the innermost integral: since 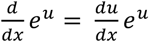, antidifferentiation gives 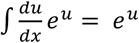. In this case, u being −*i.t*. we get

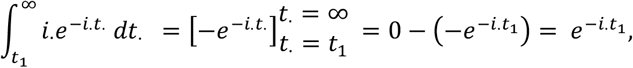

therefore

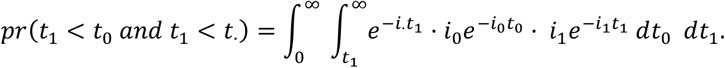

Similarly, the second integral becomes 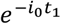, so that

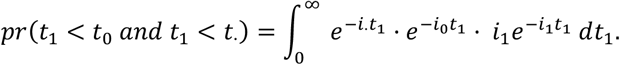

Since i_1_ is constant we can move it outside of the integral:

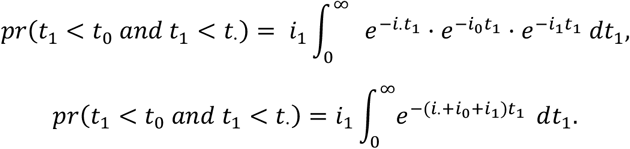

Multiplying and dividing the integral by the same constant (*i*_·_+ *i*_0_ + *i*_1_):

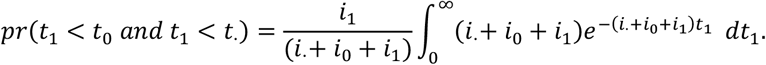

After integration we get

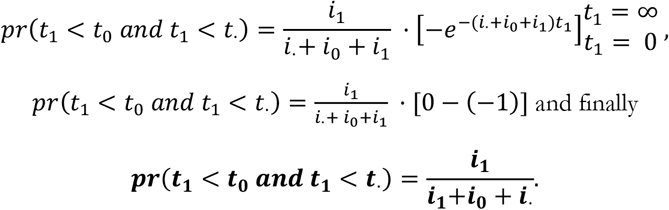

Similarly, one gets

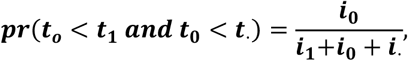

and

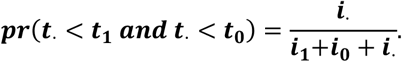

### 4 Derivation of the CF from I_E_ and I_U_ without recourse to the concept of causes

If t_E_ and t_U_, the individual occurrence times of the disease under exposure and under non exposure, respectively, are independent of each other in all individuals, and if I_E_ and I_U_, the observed disease occurrence rates, are constant over time, we have

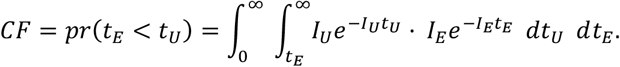

Solving the inner integral:

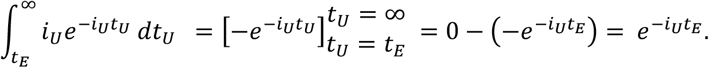

Then the outer integral then becomes

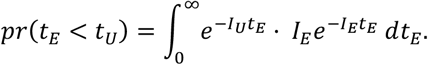

After moving the constant I_E_ outside of the integral and after multiplying and dividing by (*I*_*E*_ + *I*_*U*_) we get

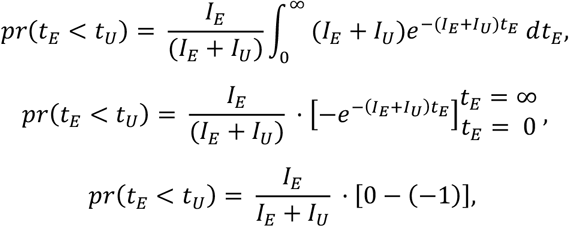

and finally

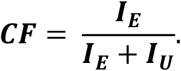

### 5 Derivation of the CF from a constant HR, with I_E_ and I_U_ constant over time

As is the case for the EF and the PF, the CF can be expressed as a function of the hazard ratio HR = I_E_/I_U_, so that I_E_ = I_U_HR. Under independence of t_E_ and t_U_ the joint probability density function is

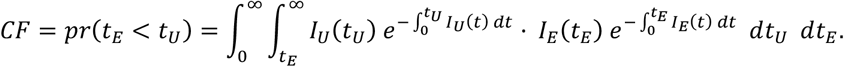

Under constancy of rates 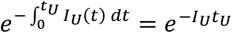and 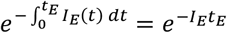 (Appendix 2), so that we now have

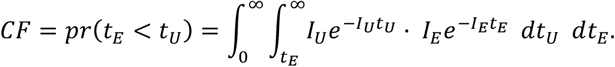

Since I_E_ = I_U_HR, this becomes

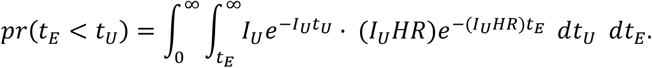

Solving the inner integral:

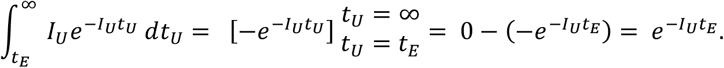

The outer integral then becomes

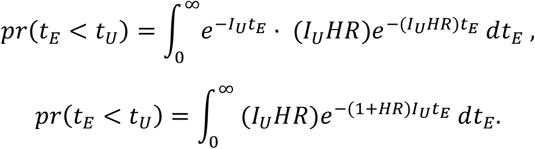

After moving the constant HR outside of the integral, and multiplying and dividing by the constant (1+HR) we get

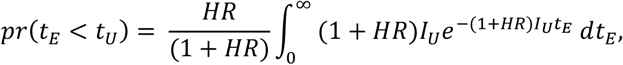

so that

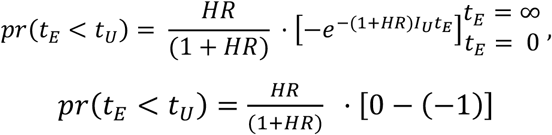

and finally

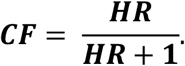

### 6 Derivation of the CF from a constant HR, with I_E_(t) and I_U_(t) as functions of time

Under the less restrictive assumption of a constant HR, even though the incidence rates are time-dependent, we have I_E_(t) = I_U_(t)HR. Under independence of t_E_ and t_U_, the first integral in Appendix 5 becomes

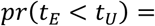

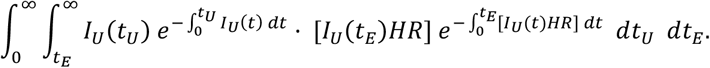

Beginning with the inner integral

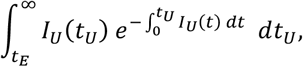

Since, by antidifferentiation,

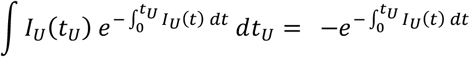

we have

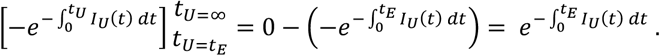

The outer integral then becomes

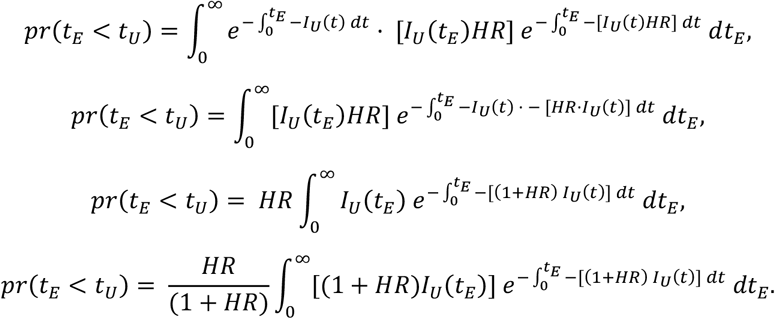

Since

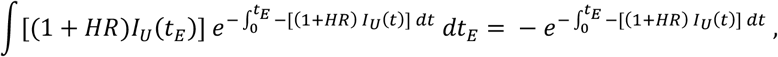

we get

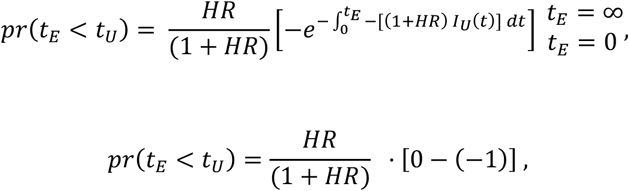

and finally

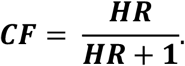

Note: This last result depends on the function I_U_(t) allowing 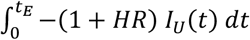 to → ∞ when *t*_*E*_ → ∞ (see Appendix 2).

### 7 SPSS syntax for estimating maxCF and minCF from distributions of disease occurrence times under exposure and non-exposure, with examples

**Figure 1.**
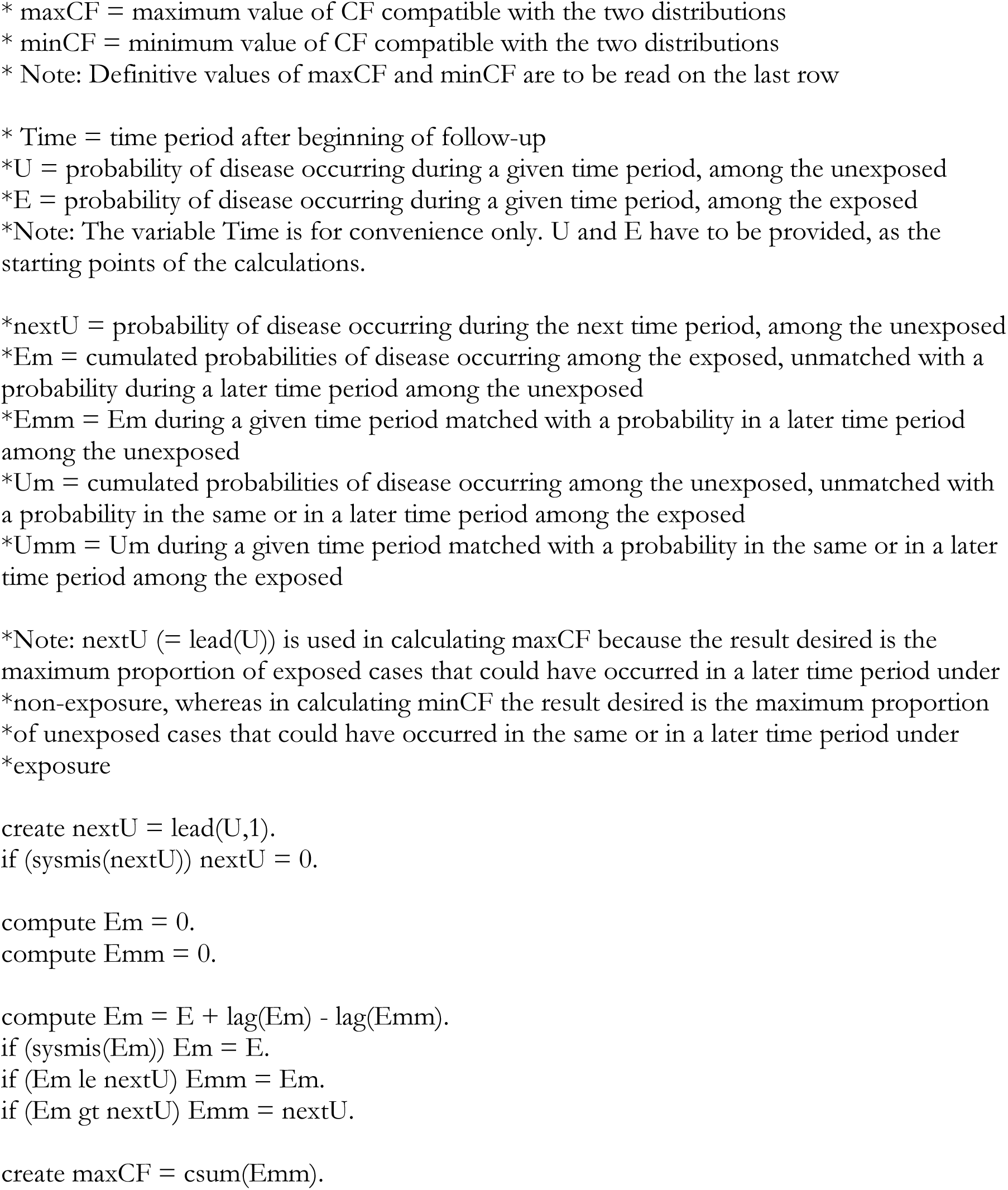
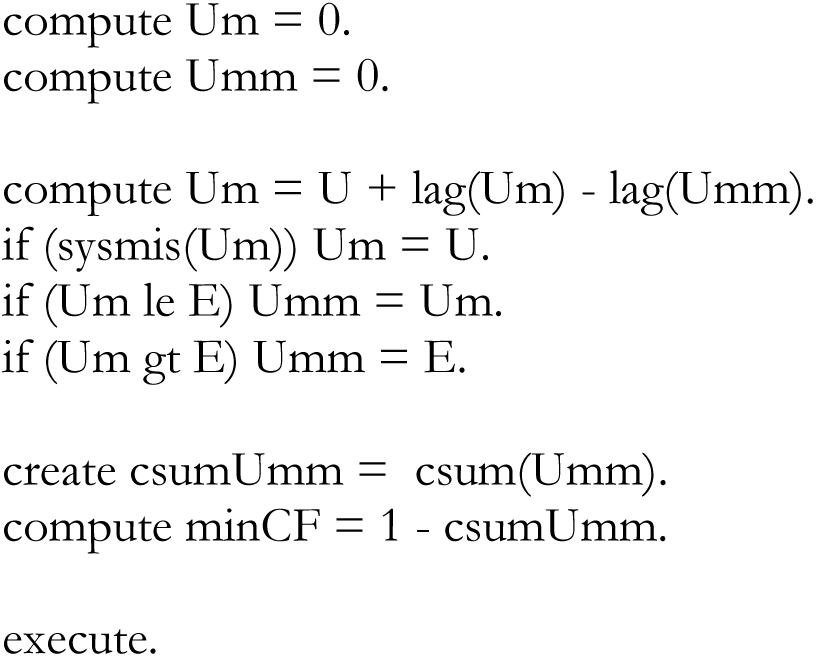
SPSS syntax.

**Table 1.** Two identical distributions, under exposure and non-exposure; 0≤CF≤0,88. The distributions are compatible with a null effect of the exposure, but also with strong preventive and causative effects.

| Time | E | U | nextU | Em | Emm | maxCF | Um | Umm | csumUmm | minCF |
| --- | --- | --- | --- | --- | --- | --- | --- | --- | --- | --- |
| 1 | 0 | 0 | 0,01 | 0 | 0 | 0 | 0 | 0 | 0 | 1 |
| 2 | 0,01 | 0,01 | 0,02 | 0,01 | 0,01 | 0,01 | 0,01 | 0,01 | 0,01 | 0,99 |
| 3 | 0,02 | 0,02 | 0,03 | 0,02 | 0,02 | 0,03 | 0,02 | 0,02 | 0,03 | 0,97 |
| 4 | 0,03 | 0,03 | 0,04 | 0,03 | 0,03 | 0,06 | 0,03 | 0,03 | 0,06 | 0,94 |
| 5 | 0,04 | 0,04 | 0,05 | 0,04 | 0,04 | 0,1 | 0,04 | 0,04 | 0,1 | 0,9 |
| 6 | 0,05 | 0,05 | 0,06 | 0,05 | 0,05 | 0,15 | 0,05 | 0,05 | 0,15 | 0,85 |
| 7 | 0,06 | 0,06 | 0,08 | 0,06 | 0,06 | 0,21 | 0,06 | 0,06 | 0,21 | 0,79 |
| 8 | 0,08 | 0,08 | 0,1 | 0,08 | 0,08 | 0,29 | 0,08 | 0,08 | 0,29 | 0,71 |
| 9 | 0,1 | 0,1 | 0,11 | 0,1 | 0,1 | 0,39 | 0,1 | 0,1 | 0,39 | 0,61 |
| 10 | 0,11 | 0,11 | 0,12 | 0,11 | 0,11 | 0,5 | 0,11 | 0,11 | 0,5 | 0,5 |
| 11 | 0,12 | 0,12 | 0,09 | 0,12 | 0,09 | 0,59 | 0,12 | 0,12 | 0,62 | 0,38 |
| 12 | 0,09 | 0,09 | 0,08 | 0,12 | 0,08 | 0,67 | 0,09 | 0,09 | 0,71 | 0,29 |
| 13 | 0,08 | 0,08 | 0,06 | 0,12 | 0,06 | 0,73 | 0,08 | 0,08 | 0,79 | 0,21 |
| 14 | 0,06 | 0,06 | 0,05 | 0,12 | 0,05 | 0,78 | 0,06 | 0,06 | 0,85 | 0,15 |
| 15 | 0,05 | 0,05 | 0,04 | 0,12 | 0,04 | 0,82 | 0,05 | 0,05 | 0,9 | 0,1 |
| 16 | 0,04 | 0,04 | 0,03 | 0,12 | 0,03 | 0,85 | 0,04 | 0,04 | 0,94 | 0,06 |
| 17 | 0,03 | 0,03 | 0,02 | 0,12 | 0,02 | 0,87 | 0,03 | 0,03 | 0,97 | 0,03 |
| 18 | 0,02 | 0,02 | 0,01 | 0,12 | 0,01 | 0,88 | 0,02 | 0,02 | 0,99 | 0,01 |
| 19 | 0,01 | 0,01 | 0 | 0,12 | 0 | 0,88 | 0,01 | 0,01 | 1 | 0 |
| 20 | 0 | 0 | 0 | 0,12 | 0 | 0,88 | 0 | 0 | 1 | 0 |

**Table 2.** Strong causative effect; 0,56≤CF≤1. The distributions are only compatible with a causative effect of the exposure.

| Time | E | U | nextU | Em | Emm | maxCF | Um | Umm | csumUmm | minCF |
| --- | --- | --- | --- | --- | --- | --- | --- | --- | --- | --- |
| 1 | 0 | 0 | 0 | 0 | 0 | 0 | 0 | 0 | 0 | 1 |
| 2 | 0,01 | 0 | 0 | 0,01 | 0 | 0 | 0 | 0 | 0 | 1 |
| 3 | 0,01 | 0 | 0 | 0,02 | 0 | 0 | 0 | 0 | 0 | 1 |
| 4 | 0,05 | 0 | 0,01 | 0,07 | 0,01 | 0,01 | 0 | 0 | 0 | 1 |
| 5 | 0,1 | 0,01 | 0,02 | 0,16 | 0,02 | 0,03 | 0,01 | 0,01 | 0,01 | 0,99 |
| 6 | 0,12 | 0,02 | 0,02 | 0,26 | 0,02 | 0,05 | 0,02 | 0,02 | 0,03 | 0,97 |
| 7 | 0,2 | 0,02 | 0,05 | 0,44 | 0,05 | 0,1 | 0,02 | 0,02 | 0,05 | 0,95 |
| 8 | 0,15 | 0,05 | 0,08 | 0,54 | 0,08 | 0,18 | 0,05 | 0,05 | 0,1 | 0,9 |
| 9 | 0,1 | 0,08 | 0,1 | 0,56 | 0,1 | 0,28 | 0,08 | 0,08 | 0,18 | 0,82 |
| 10 | 0,09 | 0,1 | 0,12 | 0,55 | 0,12 | 0,4 | 0,1 | 0,09 | 0,27 | 0,73 |
| 11 | 0,05 | 0,12 | 0,18 | 0,48 | 0,18 | 0,58 | 0,13 | 0,05 | 0,32 | 0,68 |
| 12 | 0,04 | 0,18 | 0,15 | 0,34 | 0,15 | 0,73 | 0,26 | 0,04 | 0,36 | 0,64 |
| 13 | 0,03 | 0,15 | 0,1 | 0,22 | 0,1 | 0,83 | 0,37 | 0,03 | 0,39 | 0,61 |
| 14 | 0,02 | 0,1 | 0,07 | 0,14 | 0,07 | 0,9 | 0,44 | 0,02 | 0,41 | 0,59 |
| 15 | 0,01 | 0,07 | 0,04 | 0,08 | 0,04 | 0,94 | 0,49 | 0,01 | 0,42 | 0,58 |
| 16 | 0,01 | 0,04 | 0,03 | 0,05 | 0,03 | 0,97 | 0,52 | 0,01 | 0,43 | 0,57 |
| 17 | 0,01 | 0,03 | 0,02 | 0,03 | 0,02 | 0,99 | 0,54 | 0,01 | 0,44 | 0,56 |
| 18 | 0 | 0,02 | 0,01 | 0,01 | 0,01 | 1 | 0,55 | 0 | 0,44 | 0,56 |
| 19 | 0 | 0,01 | 0 | 0 | 0 | 1 | 0,56 | 0 | 0,44 | 0,56 |
| 20 | 0 | 0 | 0 | 0 | 0 | 1 | 0,56 | 0 | 0,44 | 0,56 |

**Table 3.** Weak preventive effect; 0≤CF≤0,51. The distributions are compatible with a preventive or null effect of the exposure but also with a very weak causative effect.

| Time | E | U | nextU | Em | Emm | maxCF | Um | Umm | csumUmm | minCF |
| --- | --- | --- | --- | --- | --- | --- | --- | --- | --- | --- |
| 1 | 0 | 0 | 0,01 | 0 | 0 | 0 | 0 | 0 | 0 | 1 |
| 2 | 0 | 0,01 | 0,01 | 0 | 0 | 0 | 0,01 | 0 | 0 | 1 |
| 3 | 0 | 0,01 | 0,01 | 0 | 0 | 0 | 0,02 | 0 | 0 | 1 |
| 4 | 0 | 0,01 | 0,02 | 0 | 0 | 0 | 0,03 | 0 | 0 | 1 |
| 5 | 0,01 | 0,02 | 0,05 | 0,01 | 0,01 | 0,01 | 0,05 | 0,01 | 0,01 | 0,99 |
| 6 | 0,01 | 0,05 | 0,1 | 0,01 | 0,01 | 0,02 | 0,09 | 0,01 | 0,02 | 0,98 |
| 7 | 0,03 | 0,1 | 0,12 | 0,03 | 0,03 | 0,05 | 0,18 | 0,03 | 0,05 | 0,95 |
| 8 | 0,05 | 0,12 | 0,2 | 0,05 | 0,05 | 0,1 | 0,27 | 0,05 | 0,1 | 0,9 |
| 9 | 0,08 | 0,2 | 0,15 | 0,08 | 0,08 | 0,18 | 0,42 | 0,08 | 0,18 | 0,82 |
| 10 | 0,1 | 0,15 | 0,1 | 0,1 | 0,1 | 0,28 | 0,49 | 0,1 | 0,28 | 0,72 |
| 11 | 0,12 | 0,1 | 0,09 | 0,12 | 0,09 | 0,37 | 0,49 | 0,12 | 0,4 | 0,6 |
| 12 | 0,15 | 0,09 | 0,05 | 0,18 | 0,05 | 0,42 | 0,46 | 0,15 | 0,55 | 0,45 |
| 13 | 0,18 | 0,05 | 0,04 | 0,31 | 0,04 | 0,46 | 0,36 | 0,18 | 0,73 | 0,27 |
| 14 | 0,1 | 0,04 | 0,03 | 0,37 | 0,03 | 0,49 | 0,22 | 0,1 | 0,83 | 0,17 |
| 15 | 0,07 | 0,03 | 0,02 | 0,41 | 0,02 | 0,51 | 0,15 | 0,07 | 0,9 | 0,1 |
| 16 | 0,04 | 0,02 | 0 | 0,43 | 0 | 0,51 | 0,1 | 0,04 | 0,94 | 0,06 |
| 17 | 0,03 | 0 | 0 | 0,46 | 0 | 0,51 | 0,06 | 0,03 | 0,97 | 0,03 |
| 18 | 0,02 | 0 | 0 | 0,48 | 0 | 0,51 | 0,03 | 0,02 | 0,99 | 0,01 |
| 19 | 0,01 | 0 | 0 | 0,49 | 0 | 0,51 | 0,01 | 0,01 | 1 | 0 |
| 20 | 0 | 0 | 0 | 0,49 | 0 | 0,51 | 0 | 0 | 1 | 0 |

## Author Contribution

Sole-author contribution.

## Ethics approval

The work reported in this communication is purely conceptual, uses no information on any human subject and required no ethics approval.

